# The World Mortality Dataset: Tracking excess mortality across countries during the COVID-19 pandemic

**DOI:** 10.1101/2021.01.27.21250604

**Authors:** Ariel Karlinsky, Dmitry Kobak

## Abstract

Comparing the impact of the COVID-19 pandemic between countries or across time is difficult because the reported numbers of cases and deaths can be strongly affected by testing capacity and reporting policy. Excess mortality, defined as the increase in all-cause mortality relative to the expected mortality, is widely considered as a more objective indicator of the COVID-19 death toll. However, there has been no global, frequently-updated repository of the all-cause mortality data across countries. To fill this gap, we have collected weekly, monthly, or quarterly all-cause mortality data from 94 countries and territories, openly available as the regularly-updated World Mortality Dataset. We used this dataset to compute the excess mortality in each country during the COVID-19 pandemic. We found that in several worst-affected countries (Peru, Ecuador, Bolivia, Mexico) the excess mortality was above 50% of the expected annual mortality. At the same time, in several other countries (Australia, New Zealand) mortality during the pandemic was below the usual level, presumably due to social distancing measures decreasing the non-COVID infectious mortality. Furthermore, we found that while many countries have been reporting the COVID-19 deaths very accurately, some countries have been substantially underreporting their COVID-19 deaths (e.g. Nicaragua, Russia, Uzbekistan), sometimes by two orders of magnitude (Tajikistan). Our results highlight the importance of open and rapid all-cause mortality reporting for pandemic monitoring.

## 1 Introduction

The impact of COVID-19 on a given country is usually assessed via the number of cases and the number of deaths, two statistics that have been reported daily by each country and put together into international dashboards such as the ones maintained by the World Health Organization (https://covid19.who.int) or by the Johns Hopkins University (https://coronavirus.jhu.edu) (Dong et al., 2020). However, both metrics can be heavily affected by limited testing availability and by different definitions of ‘COVID-19 death’ used by different countries (Riffe and Acosta, 2021): for example, some countries count only PCR-confirmed COVID-19 deaths, while others include suspected COVID-19 deaths as well.

Excess mortality, defined as the increase of the all-cause mortality over the mortality expected based on historic trends, has long been used to estimate the death toll of pandemics and other extreme events — from the Great Plague of London in 1665 (as described in Boka and Wainer, 2020), to the influenza epidemic in London in 1875 (Farr, 1885; Langmuir, 1976), the XX–XXI century influenza pandemics of 1918, 1957, 1968, 2009 (Murray et al., 2006; Viboud et al., 2005, 2016; Simonsen et al., 2013) as well as seasonal influenza epidemics (Housworth and Langmuir, 1974), and more recently e.g. Hurricane Maria in Puerto-Rico in 2016 (Milken Institute, 2018). Even though the excess mortality does not exactly equal the mortality from COVID-19 infections, the consensus is that for many countries it is the most objective possible indicator of the COVID-19 death toll (Beaney et al., 2020; Leon et al., 2020). Excess mortality has already been used to estimate the COVID-19 impact in different countries, both in academic literature (e.g. Kontis et al., 2020; Alicandro et al., 2020; Ghafari et al., 2020; Woolf et al., 2020a,b; Weinberger et al., 2020; Blangiardo et al., 2020; Kobak, 2021; Modi et al., 2021; Islam et al., 2021; Bradshaw et al., 2021, among many others) and by major media outlets. It has also been used to compare COVID-19 impact to the impact of major influenza pandemics (Faust et al., 2020; Petersen et al., 2020).

Measuring and monitoring excess mortality across different countries requires, first and foremost, a comprehensive and regularly-updated dataset on all-cause mortality. However, there has been no single resource where such data would be collected from all over the world. The *World Mortality Dataset* presented here aims to fill this gap by combining publicly-available information on country-level mortality, culled and harmonized from various sources.

Several teams have already started to collect such data. In April 2020, EuroStat (http://ec.europa.eu/eurostat) began collecting total weekly deaths across European countries, “in order to support the policy and research efforts related to COVID-19.” At the time of writing, this dataset covers 36 European countries and also contains sub-national (NUTS2 and NUTS3 regions) data as well as data disaggregated by age groups and by sex for some countries. In May 2020, the Human Mortality Database (http://mortality.org), a joint effort by the University of California, Berkeley, and Max Planck Institute for Demographic Research (Barbieri et al., 2015), started compiling the *Short Term Mortality Fluctuations* (STMF) dataset (STMF, 2021; Németh et al., 2021; Islam et al., 2021). This dataset consists of weekly data, disaggregated by five age groups and by sex, and currently contains 35 countries with 2020 data. STMF only includes countries with complete high quality vital registration data in all age groups. Both datasets are regularly updated and have considerable overlap, covering together 44 countries.

In parallel, the EuroMOMO project (https://www.euromomo.eu), existing since 2008, has been displaying weekly excess mortality in 23 European countries, but without giving access to the underlying data. Another source of data is the UNDATA initiative (http://data.un.org; search for ‘Deaths by month of death’) by the United Nations, collecting monthly mortality data across a large number of countries. However, information there is updated very slowly, with January–June 2020 data currently available for only four countries.

Media outlets such as the *Financial Times, The Economist*, the *New York Times*, and the *Wall Street Journal* have been compiling and openly sharing their own datasets in order to report on the all-cause mortality in 2020. However, these datasets are infrequently updated and their future is unclear. For example, the *New York Times* announced in early 2021 that they would stop tracking excess deaths due to staffing changes.

Here, we present the *World Mortality Dataset* that aims to provide regularly-updated all-cause mortality numbers from all over the world. The dataset is openly available at https://github.com/akarlinsky/world_mortality and is updated almost daily. Our dataset builds upon the EuroStat and the STMF datasets, adding 50 additional countries — many more than any previous media or academic effort. At the time of writing, our dataset comprises 94 countries and territories. After the initial release of our manuscript, the dataset has been incorporated into the excess mortality trackers by *Our World in Data* (Giattino et al., 2020), *The Economist*, and the *Financial Times*. While not all countries provide equally detailed and reliable data, we believe that information from all 94 countries is reliable enough to allow computation of excess mortality (see Discussion).

Our analysis (updated almost daily at https://github.com/dkobak/excess-mortality) showed statistically significant positive excess mortality in 68 out of 94 countries. Moreover, it suggests that the true COVID-19 death toll in several countries is over an order of magnitude larger than the official COVID-19 death count.

## 2 Methods

### 2.1 World Mortality Dataset

For all countries that are not covered by EuroStat or STMF, we aimed to collect weekly, monthly, or quarterly all-cause mortality data from their National Statistics Offices (NSOs), Population Registries, Ministries of Health, Ministries of Public Health, etc., collectively referred to here as ‘NSOs’.

Our strategy was to search for mortality numbers for every country on their NSO’s website. The data may be present in the form of a spreadsheet, a table generator, a periodical bulletin, a press release, a figure that required digitizing, etc. If we were unable to locate such data, we contacted the NSO via email, a contact form on their website, or on social media, asking them if they have weekly, monthly, or quarterly data on all-cause mortality for 2020.

Responses from NSOs have varied substantially. Some have provided us with the requested information, some replied that no such data were available, some did not respond at all. For many countries, the email addresses did not work and returned an error message. Here are some representative examples of the declining responses: “Due to the federal organization of our country, we don’t have 2020 mortality data available yet” (Argentina); “We are sorry to inform you that we do not have the data you requested” (China); “These are not available” (India); “Unfortunately we don’t have this data. Currently, we only have the number of people dying from traffic accidents (by month)” (Vietnam); “Unfortunately, we do not have a mechanism in place at the moment to capture routine mortality data in-country nation-wide […] As you may also be aware, death or mortality registration or reporting is yet a huge challenge in developing countries […]” (Liberia).

We included the data from 2015 onwards into our dataset if it satisfied the following inclusion criteria: (1) data were in weekly, monthly or quarterly format (we preferred weekly data whenever available); (2) data existed at least until June 2020; (3) there were data for at least one entire year before 2020 (or a forecast for 2020, see below). At the time of writing, our dataset comprises 94 countries and territories (Figure 1).

**Figure 1:**
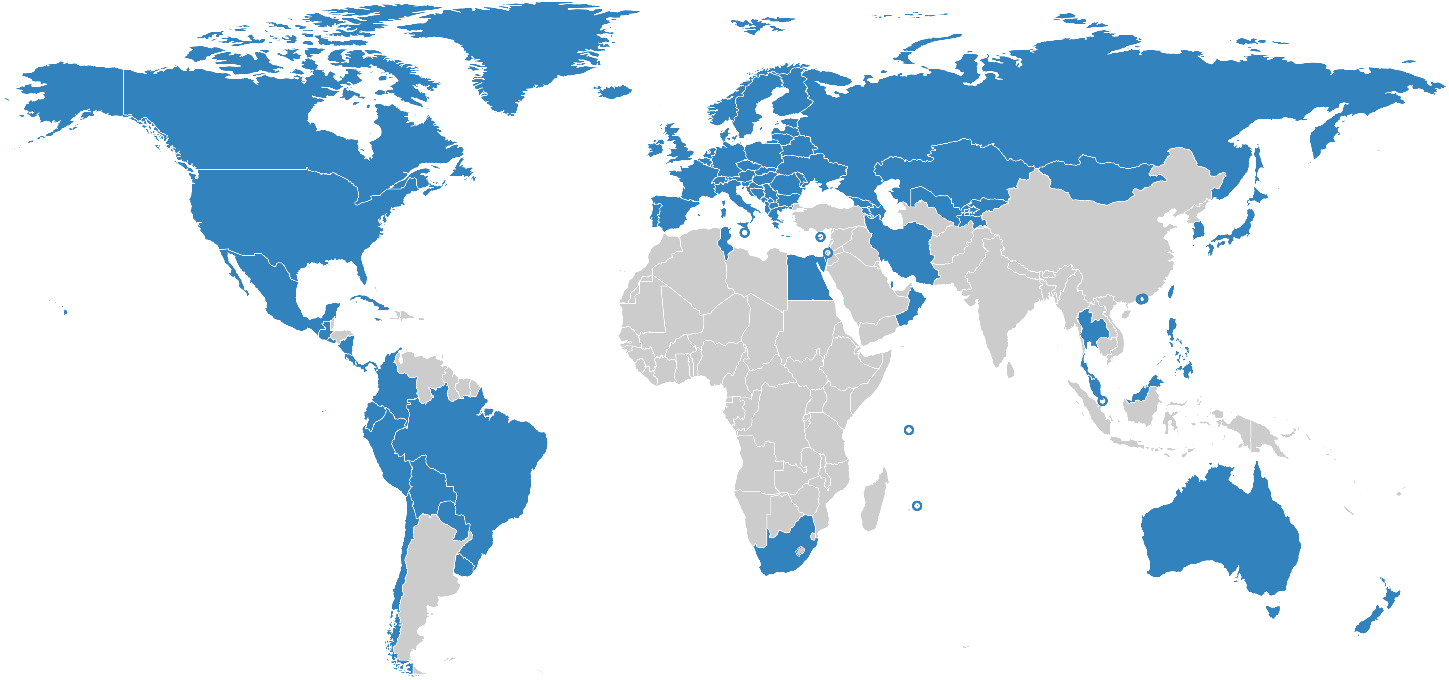
Countries in World Mortality Dataset are shown in blue. Cyprus, Hong Kong, Israel, Macao, Malta, Mauritius and Singapore are shown as circles due to their small geographical size.

For the weekly data, we preferred ISO weeks whenever possible (for Peru, Sweden, Ecuador, and Guatemala we converted daily data into ISO weeks). For Liechtenstein and Taiwan we preferred monthly format over weekly data from STMF/EuroStat, because weekly data were very noisy or less up-to-date. The data for Iran are available in quarterly format, where quarters start on December 21, March 21, June 21, and September 21 (Solar Hijri seasons). We treated the season starting on December 21 as the first data point for the following year.

Unlike STMF (Islam et al., 2021), we only collected country-level data, without age or gender stratification, since for most countries this information was not available. In some cases, we had to combine several data sources, for example taking the 2015–2018 monthly data from UNDATA and 2019–2020 monthly data from a country’s NSO. For three countries (El Salvador, Gibraltar, Nicaragua) some of the values were taken from media reports which in turn obtained them from the respective NSOs. A detailed description of all data sources for each country can be found at https://github.com/akarlinsky/world_mortality.

In this manuscript, we treat Taiwan, Hong Kong, and Macao as separate countries. They release monthly all-cause mortality data, whereas China does not. We also treat Gibraltar, Greenland, and Transnistria as separate territories as the United Kingdom, Denmark, and Moldova do not report these deaths in their figures.

### 2.2 Data limitations and caveats

The data in our dataset come with several important caveats.

First, the 2020–2021 data are often preliminary and subject to backward revisions. The more recent the data point, the more incomplete it usually is. Some countries only publish complete data (with a substantial delay) while others release very early and incomplete data as well. We excluded the most recent data points whenever there was an indication that the data were substantially incomplete. For the United States, we used the ‘weighted’ mortality counts from the Centers for Disease Control and Prevention (CDC) that account for undercount in recent weeks, instead of the STMF data.

Second, the completeness and reliability of all-cause mortality data varies by country. According to the United Nations Demographic Yearbook (UNSD, 2019), 74 of the countries in our dataset have a death registration coverage rate of 90% and above. In the remaining 20 countries, coverage is either estimated to be below 90% (e.g. Peru, Ecuador, Bolivia) or no estimate exists at all (Kosovo, Taiwan, Transnistria). However, some of the available coverage estimates are outdated. For example, the estimate for Bolivia is from 2000. The estimate for Peru is from 2015, i.e. before the SINADEF reform in 2016 (Vargas-Herrera et al., 2018) which has likely substantially improved the coverage. For Taiwan, Human Mortality Database estimates that the data are over 99% complete. At the same time, note that the coverage estimates refer to the finalized data so preliminary 2020–21 data may be less complete, as explained above. It is also possible that COVID-19 pandemic could have affected the quality of the vital registration.

Third, whereas we preferred data by date of death, the data from many countries is only available by the date of registration. Most weekly data is by the date of death (one notable exception is United Kingdom), but monthly data are often by the date of registration. Whenever the data are organized by the date of registration, it will show spurious drops in weeks or months with public holidays (e.g. last week of August and last week of December in the United Kingdom) or during national lockdowns (e.g. April 2020 in Kyrgyzstan, Kazakhstan, and Panama).

Fourth, we aimed to collect information from all countries from 2015 onward, yet currently we only have later data for four countries: Chile (2016), Germany (2016), Transnistria (2016), Peru (2017). One other country, South Africa, did not release any pre-2020 data at all, but instead published a forecast for 2020 based on the prior data (Bradshaw et al., 2021). We included this published forecast into the dataset as year 0.

Fifth, for most countries, the data are provided as-is, but for two countries (Brazil and Sweden) we performed some processing to assure consistency across years, resulting in non-integer values. In Brazil there are two mortality monitoring system: ‘Registro Civil’ (RC) and ‘Sistema de Informaçāo sobre Mortalidade’ (SIM). RC is more up-to-date, whereas SIM is more complete. We used the SIM data up until October 2020 and RC data afterwards, multiplied by the ratio between total January–October 2020 deaths in SIM and in RC (1.08). Sweden has a substantial number of deaths (2.9% of all deaths in 2019; 2.7% in 2020) reported with an ‘unknown’ week. However, ∼95% of these have a known month of death. In order to account for this, we redistributed deaths with known month but unknown week uniformly across weeks of the respective month, and similarly redistributed the remaining deaths with known year but unknown month.

Despite the caveats and limitations listed above, all our data are self-consistent: the baseline mortality that we predict for 2020 agrees very well with the pre-COVID early 2020 mortality in all cases. Note that our projection for 2020 uses a linear trend (see Section 2.3) and so can implicitly account for improvements in death registration over the recent years. We therefore believe that for countries with incomplete death registration coverage, our excess mortality estimates provide a lower bound to the true excess mortality.

### 2.3 Excess mortality

In order to estimate the excess mortality, we first estimated the expected, or baseline, mortality for 2020 using the historical data from 2015–2019 (or as many years from this interval as were available; see above). We fitted the following regression model separately for each country:

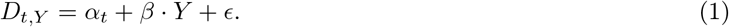

Here *D*_*t,Y*_ is the number of deaths observed on week (or month, or quarter) *t* in year *Y, β* is a linear slope across years, *α*_*t*_ are separate intercepts (fixed effects) for each week (month/quarter), and *ϵ* ∼ 𝒩(0, *σ*^2^) is Gaussian noise. This model can capture both seasonal variation in mortality and a yearly trend over recent years due to changing population structure or socio-economic factors.

As an example, using monthly death data from Russia (*R*^2^ = 0.72, *F* = 10.2), we obtained 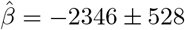 (± standard error), meaning that each year the number of monthly deaths decreases on average by ∼2300, and so the predicted monthly deaths for 2020 are ∼7000 lower than the 2015–2019 average. In contrast, using weekly data from the United States (*R*^2^ = 0.89, *F* = 31.7), we obtained 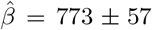, meaning that each year the number of weekly deaths increases on average by ∼800. In these two cases, as well as many other ones, the yearly trend was strong and statistically significant, and using the average 2015–2019 data as baseline, as is sometimes done, would therefore not be appropriate.

We took the model prediction for 2020 as the baseline for excess mortality calculations:

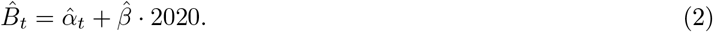

For the countries with weekly data, the model was fit using weeks 1–52, as the week 53 only happens in rare years (including 2020). The baseline for week 53 was then taken as equal to the value obtained for week 52. We took the same baseline for 2021 as for 2020, to avoid further extrapolation.

The excess mortality in each week (or month, or quarter) was defined as the difference between the actually observed death number and the baseline prediction. Note that the excess mortality can be negative, whenever the observed number of deaths is below the baseline. We summed the excess mortality estimates across all weeks starting from March 2020 (week 10; for monthly data, we started summation from March 2020; for quarterly data, from the beginning of 2020). This yields the final estimate of the excess mortality:

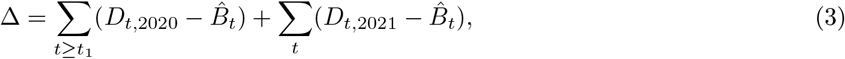

where *t*_1_ denotes the beginning of summation in 2020.

We computed the variance Var[Δ] of our estimator Δ as follows. Let **X** be the predictor matrix in the regression, **y** be the response matrix, 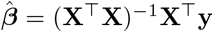 be the vector of estimated regression coefficients, and 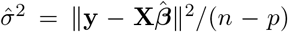 be the unbiased estimate of the noise variance, where *n* is the sample size and *p* is the number of predictors. Then 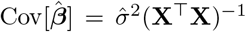 is the covariance matrix of 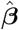 and 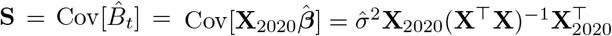 is the covariance matrix of the predicted baseline values 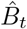 where **X**_2020_ is the predictor matrix for the entire 2020. We introduce vector **w** with elements *w*_*t*_ of length equal to the number of rows in **X**_2020_, set all elements before *t*_1_ to zero, all elements starting from *t*_1_ to 1, and increase by 1 all elements corresponding to the existing 2021 data. Then the variance of Δ is given by

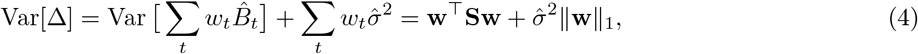

where the first term corresponds to the uncertainty of 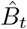 and the second term corresponds to the additive Gaussian noise that 2020–2021 observations would have had on top of *B*_*t*_ without the pandemic event. We took the square root of Var[Δ] as the standard error of Δ. Whenever the fraction 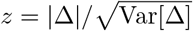 was below 2, we considered the excess mortality for that country to be not significantly different from zero. Note that we could not estimate the uncertainty for South Africa because raw historical data were not available (see above).

There exist more elaborate statistical approaches for estimating the baseline (and thus the excess) mortality, for example modeling the seasonal variation using periodic splines or Fourier harmonics, or controlling for the time-varying population size and age structure, or using a Poisson model (Farrington et al., 1996; Noufaily et al., 2013), etc. We believe that our method achieves the compromise between flexibility and simplicity: it is the simplest approach that captures both the seasonal variation and the yearly trend, and is far more transparent than more elaborate methods. Note that our uncertainty estimation assumes iid noise in Eq. 1. In reality, the noise may be temporally or spatially autocorrelated, which would affect the variance of 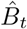.

Past (2015–2019) influenza outbreaks contributed to the estimation of the baseline 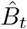. As a consequence, our baseline captures the expected mortality without the COVID-19 pandemic, but in the presence of usual seasonal influenza. This differs from the approach taken by EuroMomo as well as by some studies of excess mortality due to influenza pandemics (Viboud et al., 2005, 2016; Simonsen et al., 2013), where the baseline is constructed in a way that weighs down previous influenza outbreaks so that each new outbreak would result in positive excess mortality. A parallel work on COVID-19 excess mortality based on the STMF dataset (Islam et al., 2021) also used that approach, which explains some of the differences between our estimates.

Apart from the raw number of excess deaths, we report the number of excess deaths per 100,000 population (see below), and the number of excess deaths relative to the baseline annual deaths. For one country (El Salvador), the baseline data did not cover the entire year. There we estimated the baseline annual number of deaths as the baseline number for the existing period (January–August) times the ratio of the full year length to the length of the existing period (i.e. 12/8).

### 2.4 COVID-unrelated causes of excess mortality

We subtracted 4,000 from the excess mortality estimates for Armenia and Azerbaijan to account for the 2020 Nagorno-Karabakh war. By official counts, it cost ∼3,400 lives in Armenia and ∼2,800 in Azerbaijan (Welt and Bowen, 2021), but we took 4000 deaths in each country to obtain a conservative estimate of COVID-related excess mortality. To the best of our knowledge, no other armed conflict in 2020–2021 resulted in more than 100 casualties in countries included in our dataset.

Another correction was done for Belgium, Netherlands, France, Luxembourg, and Germany, where our data show a peak of excess deaths in August 2020, not associated with COVID-19 (see below and figure 4) and likely corresponding to a heat wave (Fouillet et al., 2006, 2008; Flynn et al., 2005). We excluded weeks 32–34 from the excess mortality calculation in these five countries. This decreased the excess mortality estimates for these countries by 1,500, 660, 1,600, 35, and 3,700, respectively.

The EM-DAT database of natural disasters (https://www.emdat.be) lists only the following four natural disasters with over 200 fatalities in 2020–2021 in countries included into our dataset: the August heat wave in Belgium, France, and the Netherlands, and a sequence of heat waves in the United Kingdom in June–August 2020 (2,500 casualties). We do not see clear peaks in our data associated with these heat waves, possibly because our United Kingdom data is by the date of registration and not by the date of death. We have therefore chosen to not adjust our excess mortality estimate for the United Kingdom.

Note that other countries may also have experienced non-COVID-related events leading to excess mortality. However, as these events are not included in the EM-DAT database, we assume that their effect would be small in comparison to the effect of COVID-19. For example, Russian data suggest ∼10,000 excess deaths from a heat wave in July 2020 in Ural and East Siberia (Kobak, 2021). We do not correct for it in this work as it is difficult to separate July 2020 excess deaths into those due to COVID and those due to the heat wave, based on the Russian country-level data alone, and we are not aware of any reliable published estimates. Importantly, 10,000 is only a small fraction of the total number of excess deaths in Russia. Another case is the February 2021 power crisis in Texas, USA that has been estimated by *BuzzFeed News* to have yielded ∼700 excess deaths (Aldhous et al., 2021). Again, this number is small compared to the total number of excess deaths in the United States.

### 2.5 Official COVID mortality

We took the officially reported COVID-19 death counts from the World Health Organization (WHO) dataset (https://covid19.who.int). To find the number of officially reported COVID-19 deaths at the time corresponding to our excess mortality estimate, we assumed that all weekly data conform to the ISO 8601 standard, and took the officially reported number on the last day of the last week available in our dataset. Some countries use non-ISO weeks (e.g. starting from January 1st), but the difference is at most several days. ISO weeks are also assumed in the ‘Data until’ column in Table 1. Officially reported numbers for Hong Kong, Macao, and Taiwan, absent in the WHO dataset, were taken from the Johns Hopkins University (JHU) dataset (https://coronavirus.jhu.edu) (Dong et al., 2020) as distributed by *Our World in Data*. We manually added officially reported numbers for Transnistria (689 by the end of February 2021; taken from the Telegram channel https://t.me/novostipmrcom).

**Table 1:**
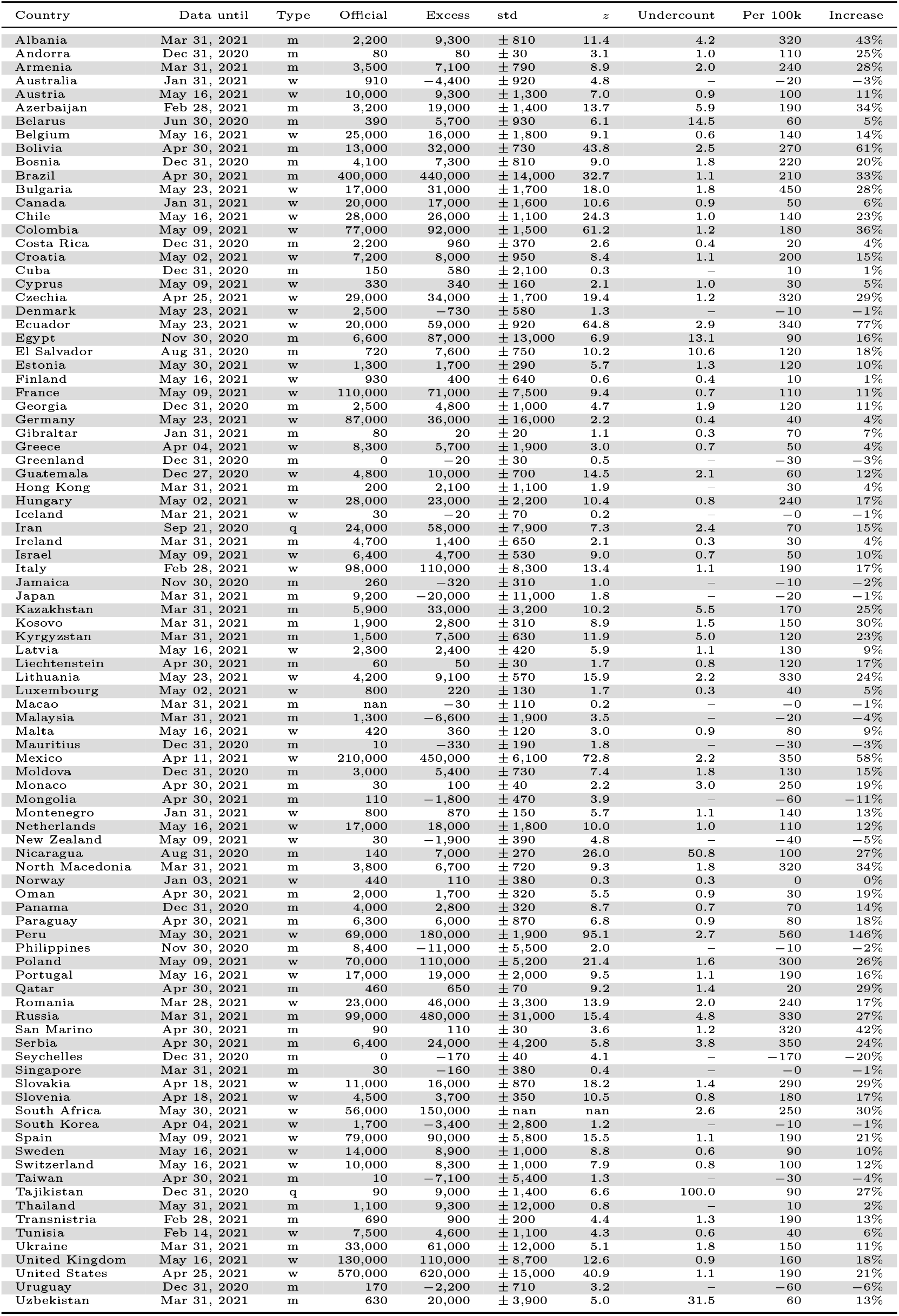
Excess mortality metrics for all countries in the dataset. Abbreviations: “w” – weekly data, “m” – monthly data, “q” – quarterly data. All numbers were rounded to two significant digits; numbers below 100 — to one significant digit. See text for the exact definitions of all reported metrics. “Official” means the official daily reported number of COVID-19 deaths. Undercount estimates are not shown for countries with negative total excess deaths and for selected countries where excess deaths were likely not related to the COVID-19 pandemic (Hong Kong, Taiwan, Cuba); see Methods.

Note that for some countries there exist different sources of official data, e.g. Russia officially reports monthly numbers of confirmed and suspected COVID deaths that are substantially larger than the daily reported numbers (Kobak, 2021). However, it is the daily reported numbers that get into the WHO and JHU dashboards, so for consistency, here we always use daily values.

We defined undercount ratio as the number of excess deaths divided by the official number of COVID-19 deaths reported by the same date. If the number of excess deaths is negative, the undercount ratio is not defined. Additionally, we chose not to show undercount ratios for countries with positive number of excess deaths where there is no evidence that excess deaths were due to COVID (Cuba, Hong Kong, Thailand). For these three countries there was no correlation between the monthly excess deaths and reported monthly numbers of COVID-19 deaths or cases, and no media reports on COVID outbreaks.

### 2.6 Population size estimates

To estimate excess deaths per 100,000 population, we obtained population size estimates for 2020 from the United Nations World Population Prospect (WPP) dataset (https://population.un.org/wpp/). The value for Russia in that dataset does not include Crimea due to its disputed status, but all Russian data of all-cause and COVID-19 mortality does include Crimea. For that reason, we used the population value of 146,748,590, provided by the Russian Federal State Statistics Service, and similarly changed the value for Ukraine to 41,762,138, provided by the Ukranian State Statistics Service. The number for Transnistria was absent in the World Population Prospect dataset, so we used the value obtained from its NSOs (465,200). Additionally, WPP reports population data for Serbia and Kosovo combined. We thus obtained population values for these countries from the World Bank Dataset (https://data.worldbank.org/indicator/SP.POP.TOTL).

Note that some of the population size estimates in the World Population Prospect dataset may be outdated or unreliable. Therefore, for some of the countries our excess death rates may be only approximate (Spoorenberg, 2020).

## 3 Results

### 3.1 Excess mortality

We collected the all-cause mortality data from 94 countries and territories into the openly-available *World Mortality Dataset*. This includes 45 countries with weekly data, 47 countries with monthly data, and 2 countries with quarterly data (Figure 1). See Methods for our data collection strategy and for some data limitations and caveats.

For each country we predicted the ‘baseline’ mortality in 2020 based on the 2015–2019 data (accounting for linear trend and seasonal variation; see Methods). We then obtained excess mortality as the difference between the actual 2020 and 2021 all-cause mortality and our baseline. For each country we computed the total excess mortality from the beginning of the COVID-19 pandemic (from March 2020) (Figure 2, Table 1). The total excess mortality was positive and significantly different from zero in 68 countries; negative and significantly different from zero in 6 countries; not significantly different from zero (*z <* 2) in 19 countries. For South Africa, there was not enough historic data available in order to assess the significance (see Methods), but the increase in mortality was very large and clearly associated with COVID-19 (Bradshaw et al., 2021).

**Figure 2:**
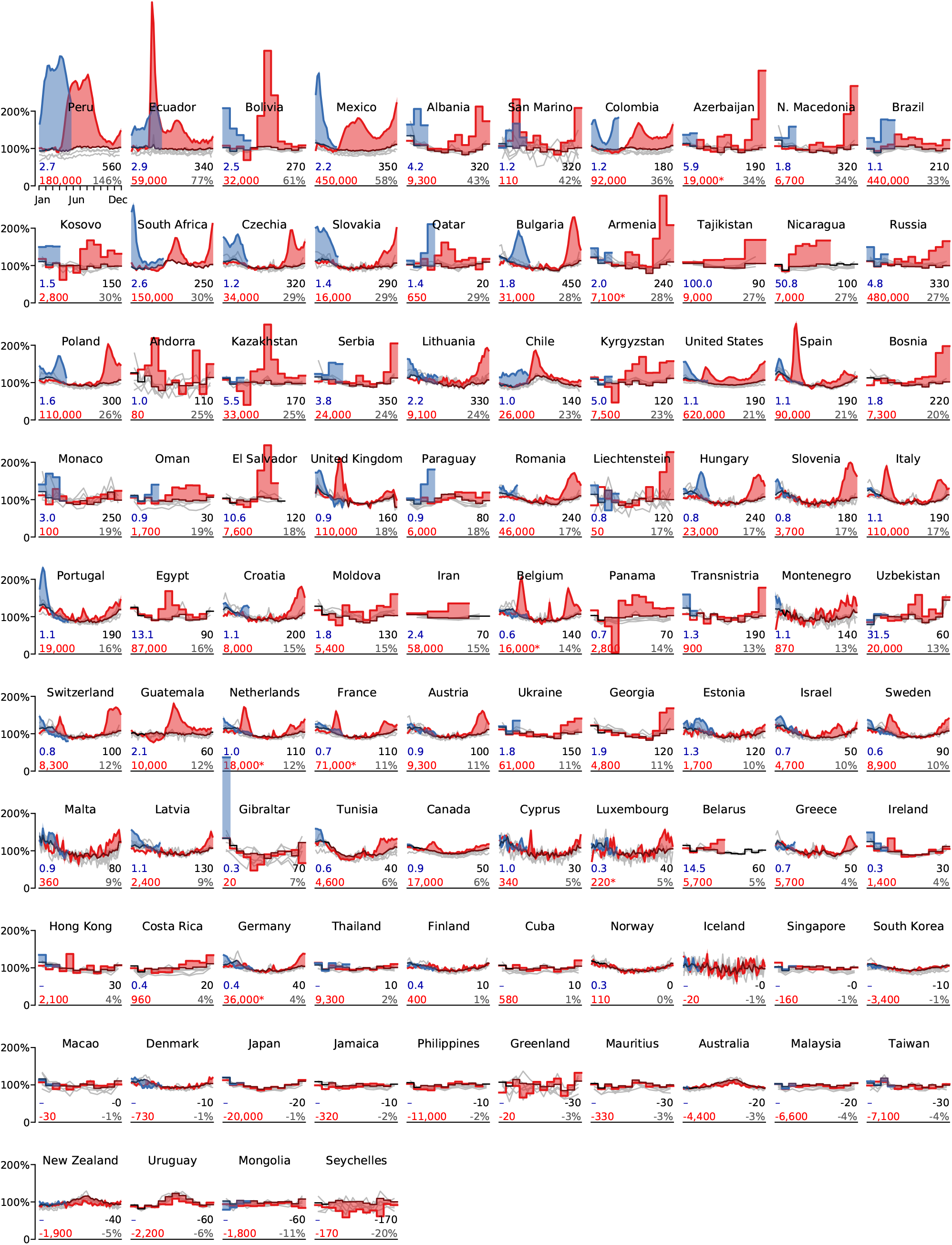
Excess mortality time series. Each subplot shows baseline mortality (black), mortality in 2015–2019 (gray), in 2020 (red) and in 2021 (blue). Excess mortality is shown in red/blue shading. The numbers in each subplot are: total excess mortality (red), excess mortality per 100,000 population (black), excess mortality as a percentage of annual baseline mortality (gray), and undercount ratio of COVID-19 deaths (blue). See text for the exact definitions. All numbers were rounded to two significant digits; numbers below 100 to one significant digit. The *y*-axis in each subplot starts at 0 and goes until 200% where 100% corresponds to the average baseline mortality. The *x*-axis covers the entire year. Asterisks mark excess mortality estimates that were downwards corrected (see Methods). Countries are sorted by the excess mortality as a percentage of annual baseline mortality (gray number). Undercount estimates are not shown for countries with negative total excess deaths and for selected countries where excess deaths were likely not related to the COVID-19 pandemic (Hong Kong, Taiwan, Cuba); see Methods.

In terms of the absolute numbers, the largest excess mortality in our dataset was observed in the United States (620,000 by April 15, 2021; all reported numbers here and below have been rounded to two significant digits), Russia (480,000 by March 31, 2021), Mexico (450,000 by April 11, 2021), and Brazil (440,000 by April 30, 2021 (Figure 3). Note that these estimates correspond to different time points as the reporting lags differ between countries (Table 1). See Figure S1 for the same analysis using the 2020 data alone.

**Figure 3:**
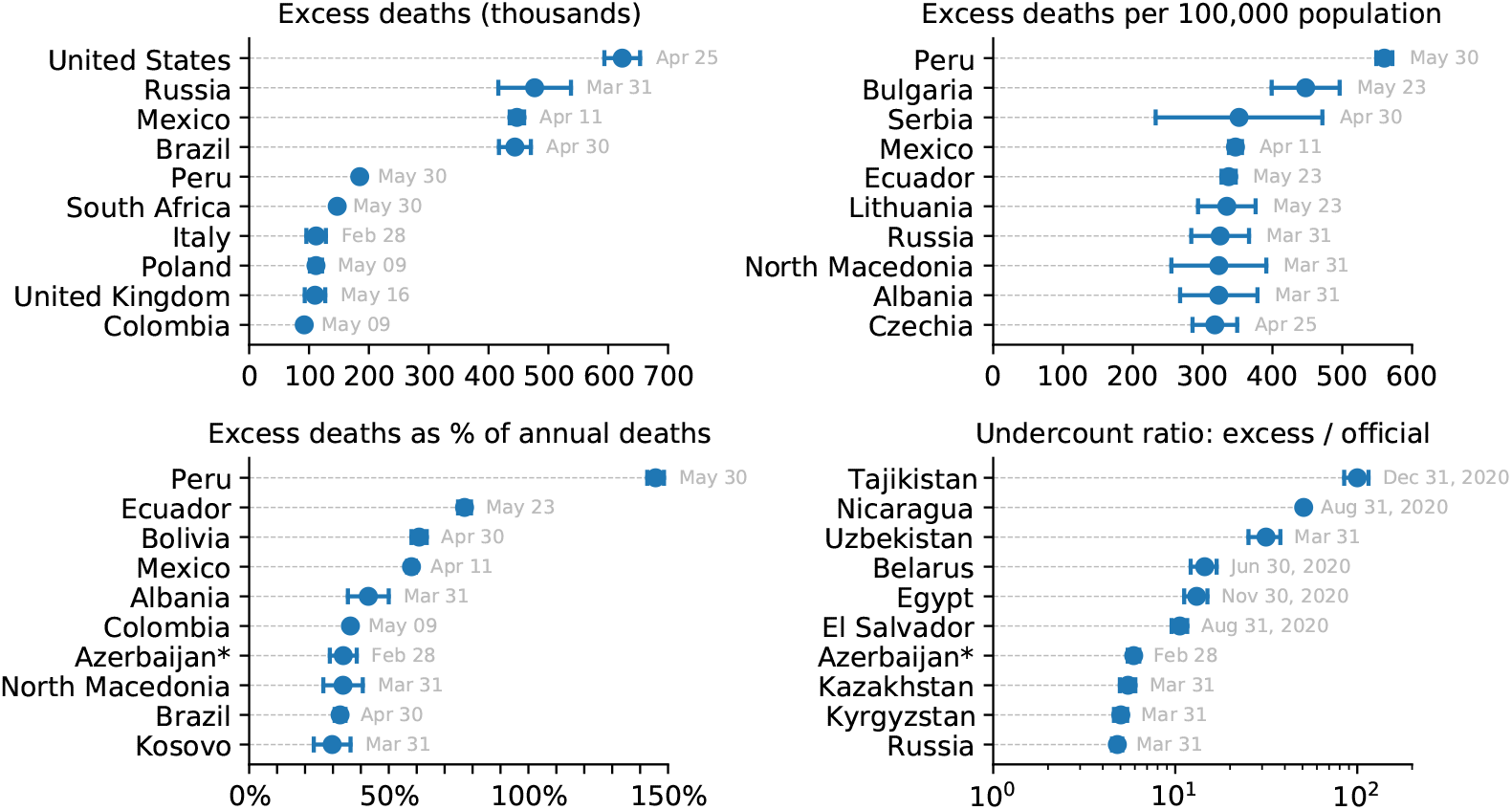
Top 10 countries in the World Mortality Dataset by various excess mortality measures. Each subplot shows the top 10 countries for each of our four excess mortality measures: total number of excess deaths; excess deaths per 100,000 population; excess deaths as a percentage of baseline annual mortality; undercount ratio (ratio of excess deaths to reported COVID-19 deaths by the same date). Error bars denote 95% confidence intervals corresponding to the uncertainty of the excess deaths estimate. Countries with population below 100,000 are not shown. Different countries have different reporting lags, so the estimates shown here correspond to different time points, as indicated. Excess mortality estimates in Armenia and Azerbaijan were downwards corrected by 4,000 to account for the war casualties (see Methods).

**Figure 4:**
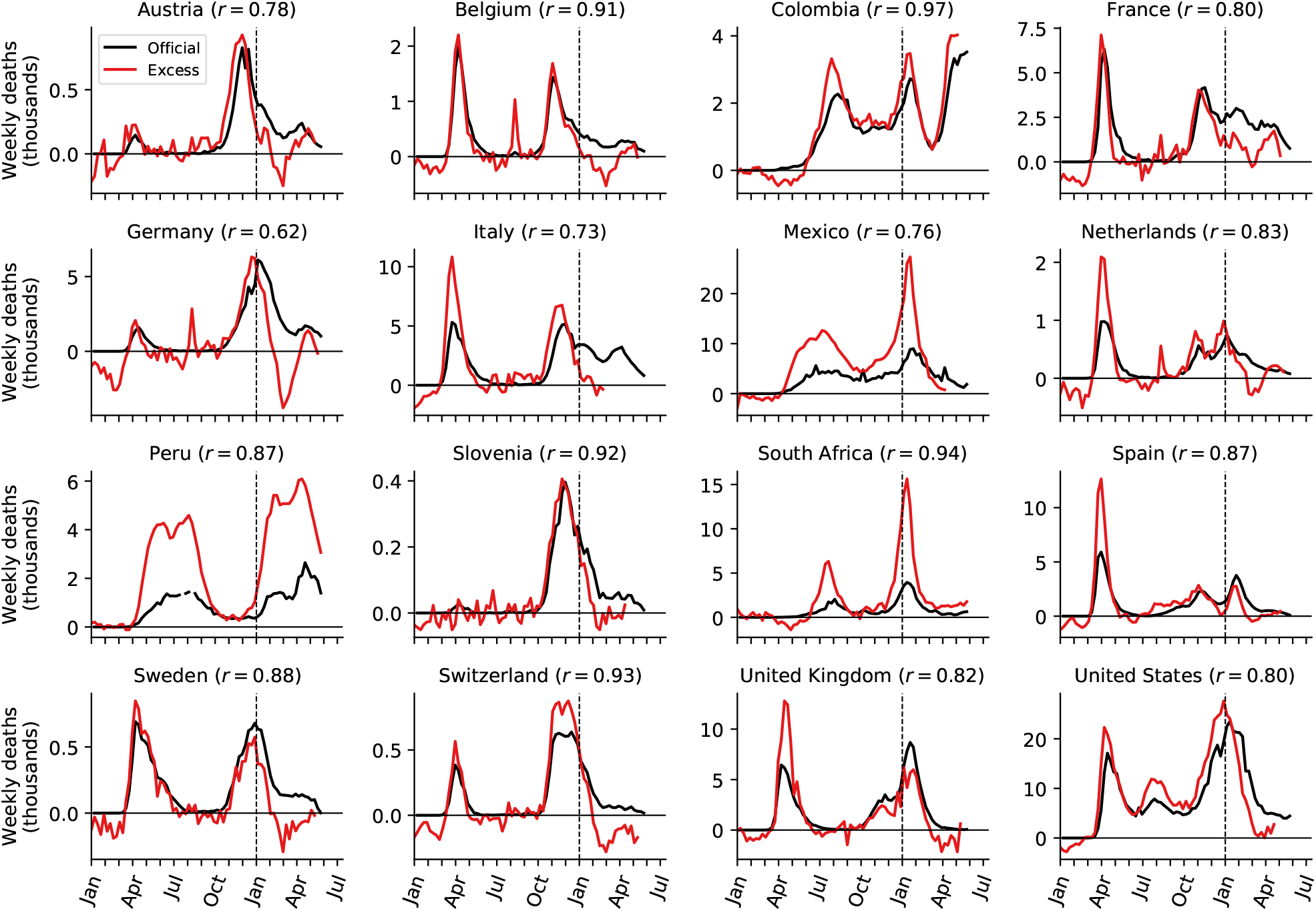
Relation between weekly excess deaths and weekly reported COVID-19 deaths. Sixteen selected countries are shown together with the Pearson correlation coefficient (*r*) between the two time series. Note the peak in excess mortality (but not in the reported COVID-19 deaths) associated with the August 2020 heat wave in Belgium, France, Germany, and Netherlands. Peru had two spikes in weekly COVID-19 deaths due to backward revisions in weeks 30 and 33 of over 5000 deaths which were removed from the plot and correlation estimation.

Some countries showed negative excess mortality, likely due to lockdown measures and social distancing decreasing the prevalence of influenza (Kung et al., 2020), as we discuss further below. For example, Australia had −4,400 excess deaths, Uruguay had −2,200 deaths, and New Zealand had −1,900 deaths. In these three cases, the decrease in mortality happened during the southern hemisphere winter season (Figure 2). Note that Uruguay had a large COVID outbreak in 2021, but we only have 2020 data available at the time of writing. The mortality decrease in Mongolia and Seychelles may also be related to the lockdown and social distancing measures but does not show clear seasonality, so may possibly also be due to other factors.

As the raw number of excess deaths can be strongly affected by the country’s population size, we normalized the excess mortality estimates by the population size (Table 1). The highest excess mortality per 100,000 inhabitants was observed in Peru (560), followed by some Latin American and East European countries: Bulgaria (450), Mexico (350), Ecuador (340), Russia (330), Albania (320), etc. (Figure 3). Note that many countries with severe outbreaks that received wide international media attention, such as Italy, Spain, and United Kingdom, had lower values (Table 1).

The infection-fatality rate (IFR) of COVID-19 is strongly age-dependent (Levin et al., 2020; O’Driscoll et al., 2021). As the countries differ in their age structure, the expected overall IFR differs between countries. To account for the age structure, we also normalized the excess mortality estimates by the annual sum of the baseline mortality, i.e. the expected number of deaths per year without a pandemic event (Table 1). This relative increase, also known as a *P-score* (Aron and Muellbauer, 2020), was by far the highest in Latin America: Peru (146%), Ecuador (77%), Bolivia (61%), and Mexico (58%) (Figure 3). These Latin American countries have much younger populations compared to the European and North American countries, which is why the excess mortality per 100,000 inhabitants there was similar to some Eastern European countries, but the relative increase in mortality was much higher, suggesting much higher COVID-19 prevalence. That the highest relative mortality increase was observed in Peru, is in agreement with some parts of Peru showing the highest measured seroprevalence level in the world (Álvarez-Antonio et al., 2021).

### 3.2 Undercount of COVID deaths

For each country we computed the ratio of the excess mortality to the officially reported COVID-19 death count by the same date. This ratio differed very strongly between countries (Table 1). Some countries had ratio below 1, e.g. 0.7 in France and 0.6 in Belgium, where reporting of COVID deaths is known to be very accurate (Sierra et al., 2020). The likely reason is that the non-COVID mortality has decreased, mostly due to the influenza suppression (see below), leading to the excess mortality underestimating the true number of COVID deaths.

Nevertheless, many countries had ratios above 1, suggesting an undercount of COVID-19 deaths (Beaney et al., 2020). At the same time, correlation between weekly reported COVID-19 deaths and weekly excess deaths was often very high (Figure 4): e.g. in Mexico (undercount ratio 2.2) correlation was *r* = 0.76; in Peru (undercount ratio 2.7) it was *r* = 0.87; in South Africa (undercount ratio 2.6) it was *r* = 0.94. High correlations suggest that excess mortality during a COVID outbreak can be fully explained by COVID-19 mortality, even when the latter is strongly underreported. See Discussion for additional considerations.

Interestingly, in many countries the undercount ratio was not constant across time. For example, the undercount ratio in Italy, Spain, Netherlands, and United Kingdom was ∼1.5 during the first wave (Figure 4), but decreased to ∼1.0 during the second wave. This decrease of the undercount ratio may be partially due to improved COVID death reporting, and partially due to the excess mortality underestimating the true COVID mortality in winter seasons due to influenza suppression.

On the other hand, several countries showed very accurate reporting of the COVID-19 deaths with the undercount ratio being close to 1.0 (Sierra et al., 2020) from the beginning of the pandemic and up until the middle of the second wave (e.g. Austria, Belgium, France, Germany, Slovenia, Sweden; Figure 4). However, starting from December 2020 and up until March 2021 the excess deaths were underestimating the COVID-19 deaths in all these countries. The difference between the officially reported COVID-19 deaths and the excess deaths may correspond to the number of deaths typically caused by influenza and other infectious respiratory diseases in winter months. This difference (computed starting from week 40 of 2020), as a fraction of baseline annual deaths, was in the 2.3–5.9% range (Austria: 2.2%, Belgium: 6.2%, France: 4.5%, Germany: 4.2%, Slovenia: 3.5%, Sweden: 5.9%), which is in good agreement with the negative excess deaths observed in Australia, New Zealand, and Uruguay (−3.0%, −5.4%, −6.4%) during the Southern hemisphere winter months (Kung et al., 2020).

The undercount ratio for most countries was below 3.0 (Table 1), but some countries showed much larger values. We found the highest undercount ratios in Tajikistan (100), Nicaragua (51), Uzbekistan (31), Belarus (14) and Egypt (13) (Figure 3). Such large undercount ratios strongly suggest purposeful misdiagnosing or underreporting of COVID-19 deaths, as argued by Kobak (2021) for the case of Russia (undercount ratio 4.8).

## 4 Discussion

### 4.1 Summary

We presented the World Mortality Dataset — the largest international dataset of all-cause mortality, currently encompassing 94 countries. The dataset is openly available and regularly updated. We are committed to keep maintaining this dataset for the entire duration of the COVID-19 pandemic.

The coverage and reliability of the data varies across countries, and some of the countries in our dataset may possibly report incomplete mortality numbers (e.g. covering only part of the country), see caveats in Section 2.2. This would make the excess mortality estimate during the COVID-19 outbreak incomplete (as an example, Lloyd-Sherlock et al. (2021) estimate that the true excess mortality in Peru may be 30% higher than excess mortality computed here due to incomplete death registration in Peru). Importantly, the early pre-outbreak 2020 data for all countries in our dataset matched the baseline obtained from the historic 2015–2019 data, indicating that the data are self-consistent and the excess mortality estimates are not inflated. Another important caveat is that the most recent data points in many countries tend to be incomplete and can experience upwards revisions. Both factors mean that some of the excess mortality estimates reported here may be underestimations.

Some of countries in our dataset have excess death estimates available in the constantly evolving literature on excess deaths during the COVID-19 pandemic from academia, official institutions and professional associations. The largest efforts include the analysis of STMF data (Kontis et al., 2020; Islam et al., 2021) and excess mortality trackers by *The Economist* and *Financial Times*. While the analysis is similar everywhere and the estimates broadly agree, there are many possible modeling choices (the start date and the end date of the total excess computation; including or excluding influenza into the baseline; modeling trend over years or not, etc.) making all the estimates slightly different.

### 4.2 Contributions to excess mortality

Conceptually, excess mortality during the COVID-19 pandemic can be represented as the sum of several distinct factors:

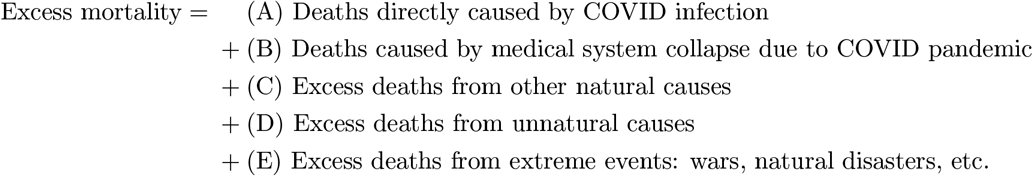

We explicitly account for factor (E) and argue that for most countries, the contribution of factors (B)–(D) is small in comparison to factor (A), in agreement with the view that excess mortality during an epidemic outbreak can be taken as a proxy for COVID-19 mortality (Beaney et al., 2020). Below we discuss each of the listed factors.

It is possible that when a country experiences a particularly strong COVID outbreak, deaths from non-COVID causes may also increase due to the medical system being overloaded — factor (B) above. Our data show that this did not happen in Belgium (undercount ratio during the first wave was close to 1.0, i.e. all excess deaths were due to COVID-19 infections), despite a ∼100% weekly increase in all-cause mortality. However, our data do not allow to say whether this factor played a role during stronger outbreaks in Latin America, with ∼200% weekly increases in all-cause mortality. Even if it did, then such collateral excess deaths can nevertheless be seen as indirect consequence of COVID-19 outbreaks.

The data suggest that the contribution of factor (C) to the excess mortality is negative. Indeed, countries that implemented stringent lockdown and social distancing measures in the absence of COVID-19 community spread, such as Australia, New Zealand, and Uruguay, showed a clear winter-season decrease in all-cause mortality, likely due to reduced influenza transmission (Kung et al., 2020). Here, using these three countries as well as some of the European data, we estimated that the influenza suppression alone can lead to a decrease of annual mortality by 3–6%. Other infectious diseases may also be suppressed by enforced social distancing. For example, in South Africa, lockdowns have noticeably decreased toddler mortality (0–4 years) (Bradshaw et al., 2021). This effect was not observed in the developed countries where toddler mortality is low (Islam et al., 2021), but could be present in other developing countries as well.

The effect of factor (D) appears to be country-specific. Traffic accident fatalities have decreased in the European Union and the Western Balkans (European Commission, 2021; Transport Community, 2021) but have increased in the United States (National Safety Council, 2021). Homicides have increased in the United States (Arthur and Asher, 2020; Faust et al., 2021) and Germany (Federal Criminal Police Office, 2021) but have decreased in Peru (Calderon-Anyosa and Kaufman, 2020), South Africa (Bradshaw et al., 2021) and France (Ministry of the Interior, 2021). Suicides have first decreased and then increased in Japan, particularly in females (Kurita et al., 2021; Tanaka and Okamoto, 2021; Black and Kutcher, 2021), have decreased in the United States (Ahmad and Anderson, 2021; Faust et al., 2021), and in a study of 21 countries were found to have decreased in half of them and remained unchanged in the rest (Pirkis et al., 2021). In the United States, deaths from drug overdoses and unintentional injuries have increased (Faust et al., 2021). However, importantly, in all cited studies the combined effect of all changes in the frequency of unnatural changes did not exceed ∼1% of the baseline annual mortality, meaning that factor (D) plays only a minor role in excess mortality.

Finally, factor (E) in 2020–2021 was mostly constrained to the Nagorno-Karabakh war between Armenia and Azerbaijan and the August 2020 heat wave in Europe, which we explicitly accounted for. We could have possibly missed some other similar events in other countries, but we believe they could only play a minor role compared to COVID-19, thanks to the absence of other major wars or natural disasters in 2020–2021.

Together, the evidence suggests that the contribution of factor (C) is negative and contribution of factor (D) is small in comparison, so we believe that lockdown and social distancing measures on their own decrease — and not increase — the death rate, at least in short-term. The contribution of factor (B), at least in developed countries, is small, and so, in the absence of wars and natural disasters, one can expect excess mortality to provide a lower bound on the true number of COVID-19 deaths. In other words, we speculate that whenever COVID deaths are counted perfectly, they should exceed the excess mortality, leading to undercount ratio below 1. This is indeed what we observed in several countries with strong COVID-19 outbreaks but accurate accounting of COVID deaths, e.g. Belgium, France, and Germany (undercount ratios 0.6, 0.7, and 0.4, respectively).

### 4.3 Conclusions and outlook

The World Mortality Dataset is open for use by researchers and policy makers from all fields. Avenues for future research include the relation between various measures of excess mortality and economic development, population structure, lockdown and social distancing measures, border controls and travel restrictions (Hale et al., 2020), properties of the health-care systems, vaccinations, institutional quality (e.g. the Democracy Index), climate, geography, population density, and many more. Conversely, future research can use excess mortality estimates to study negative social or economic impact of high COVID-19 death toll.

So far we were able to collect data from 94 nations out of ∼200, with particularly sparse coverage in Africa, Asia, and the Middle East (Figure 1). During the pandemic, many countries have sped up collection and dissemination of preliminary all-cause mortality data, yet many other countries did not, and will report 2020 information with a substantial lag, in the coming months or even years. Once released, this information will be included in the World Mortality Dataset. Unfortunately, many countries do not keep reliable vital statistics and excess mortality will remain unknown for a long time.

Summing up the excess mortality estimates across all countries in our dataset gives 3.7 million excess deaths. In contrast, summing up the official COVID-19 death counts gives 2.5 million deaths, corresponding to the global undercount ratio of 1.5. However, there is ample evidence that among the countries for which the all-cause mortality data are not available the undercount ratio is much higher (Watson et al., 2020; Djaafara et al., 2020; Watson et al., 2021; Mwananyanda et al., 2021; Besson et al., 2021). Using a statistical model to predict the excess mortality in the rest of the world based on the existing data from our dataset, *The Economist* estimated 7–13 million excess deaths worldwide (The Economist, 2021), which is 2–4 times higher than the world’s official COVID-19 death count (currently at 3.5 million).

In conclusion, the COVID-19 pandemic highlighted the great importance of reliable and up-to-date all-cause mortality data. Just as countries around the world collect and regularly report estimates of economic output such as the gross domestic product (GDP), and just as they have been reporting COVID-19 mortality, they should be reporting all-cause mortality into a comprehensive multi-national repository (Leon et al., 2020).

## Data Availability

The dataset is available at https://github.com/akarlinsky/world_mortality
The analysis code is available at https://github.com/dkobak/excess-mortality

https://github.com/dkobak/excess-mortality

## Acknowledgments

The authors would like to thank colleagues at Kohelet Policy Forum and “Midaat — for informed health”, Frédéric Fleury-Payeur, Michael Beenstock, Maxim S. Pshenichnikov, Tim Riffe, and eLife reviewers Marc Lipsitch, Ayesha Mahmud, and Lone Simonsen for their helpful comments and G. J. Andrés Uzín P., Luis Salas, Marcelo Oliveira, Otavio Ranzani, Mario Romero Zavala, Laurianne Despeghel, reporters from Eurasianet, Dmitri Tokarev, Michael Hilliard, Andrés N. Robalino, and Gilad Gaibel for their help in obtaining some of the data.

DK was supported by the Deutsche Forschungsgemeinschaft (BE5601/4-1 and the Cluster of Excellence “Machine Learning — New Perspectives for Science”, EXC 2064, project number 390727645), the Federal Ministry of Education and Research (FKZ 01GQ1601 and 01IS18039A) and the National Institute of Mental Health of the National Institutes of Health under Award Number U19MH114830. The content is solely the responsibility of the authors and does not necessarily represent the official views of the National Institutes of Health.

## Data and code availability

The World Mortality Dataset is available at https://github.com/akarlinsky/world_mortality. The analysis code is available at https://github.com/dkobak/excess-mortality. Our baseline estimates for all countries and all values shown in Table 1 are available there as CSV files.

## Author contributions

AK collected the data. AK and DK analyzed the data. AK and DK wrote the paper.

## 5 Supplementary Figures

**Figure S1:**
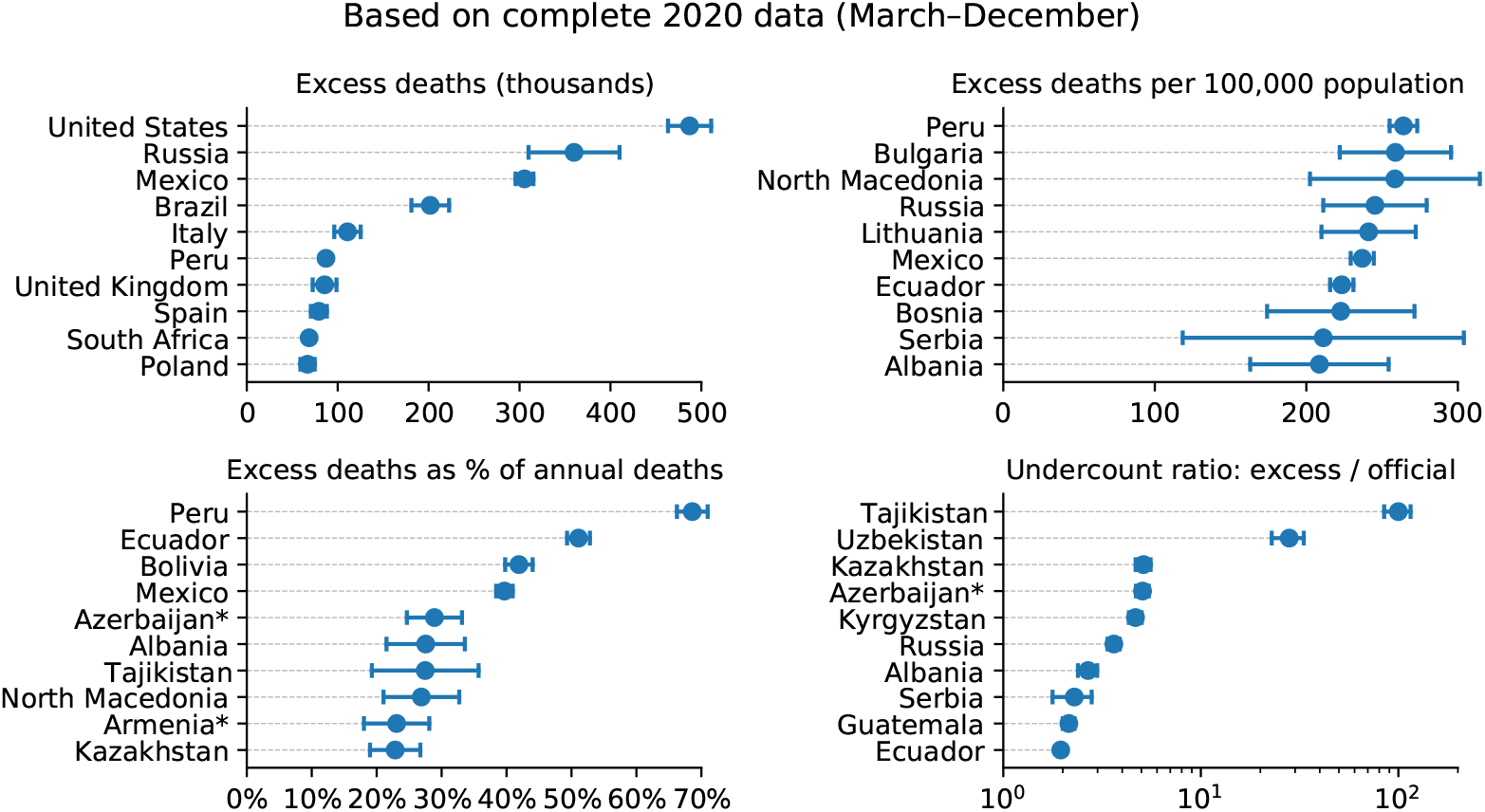
Top 10 countries in the World Mortality Dataset by various excess mortality measures by the end of 2020. Each subplot shows the top 10 countries for each of our four excess mortality measures: total number of excess deaths; excess deaths per 100,000 population; excess deaths as a percentage of baseline annual mortality; undercount ratio (ratio of excess deaths to reported COVID-19 deaths by the same date). Error bars denote 95% confidence intervals corresponding to the uncertainty of the excess deaths estimate. Countries with population below 100,000 are not shown. Excess mortality estimates in Armenia and Azerbaijan were downwards corrected by 4,000 to account for the war casualties (see Methods).

## References

Farida B. Ahmad and Robert N. Anderson. The Leading Causes of Death in the US for 2020. JAMA, 325(18): 1829–1830, 05 2021.

Peter Aldhous, Stephanie M. Lee, and Zahra Hirji. The Texas winter storm and power outages killed hundreds more people than the state says. Technical report, BuzzFeed News, 2021. URL https://www.buzzfeednews.com/article/peteraldhous/texas-winter-storm-power-outage-death-toll.

Gianfranco Alicandro, Giuseppe Remuzzi, and Carlo La Vecchia. Italy’s first wave of the COVID-19 pandemic has ended: no excess mortality in May, 2020. The Lancet, 396(10253):e27–e28, 2020.

Carlos Álvarez-Antonio, Graciela Meza-Sánchez, Carlos Calampa, Wilma Casanova, Cristiam Carey, Freddy Alava, Hugo Rodríguez-Ferrucci, and Antonio M Quispe. Seroprevalence of anti-SARS-CoV-2 antibodies in Iquitos, Peru in July and August, 2020: a population-based study. The Lancet Global Health, 2021.

Janine Aron and John Muellbauer. A pandemic primer on excess mortality statistics and their comparability across countries. Technical report, Our World in Data, 2020. URL https://ourworldindata.org/covid-excess-mortality.

Rob Arthur and Jeff Asher. What drove the historically large murder spike in 2020? Technical report, The Intercept, 2020. URL https://theintercept.com/2021/02/21/2020-murder-homicide-rate-causes/.

Magali Barbieri, John R Wilmoth, Vladimir M Shkolnikov, Dana Glei, Domantas Jasilionis, Dmitri Jdanov, Carl Boe, Timothy Riffe, Pavel Grigoriev, and Celeste Winant. Data resource profile: the human mortality database (HMD). International Journal of Epidemiology, 44(5):1549–1556, 2015.

Thomas Beaney, Jonathan M Clarke, Vageesh Jain, Amelia Kataria Golestaneh, Gemma Lyons, David Salman, and Azeem Majeed. Excess mortality: the gold standard in measuring the impact of COVID-19 worldwide? Journal of the Royal Society of Medicine, 113(9):329–334, 2020.

Emilie S Koum Besson, Andy Norris, Abdulla S Bin Ghouth, Terri Freemantle, Mervat Alhaffar, Yolanda Vazquez, Chris Reeve, Patrick J Curran, and Francesco Checchi. Excess mortality during the COVID-19 pandemic: a geospatial and statistical analysis in Aden governorate, Yemen. BMJ Global Health, 6(3):e004564, 2021.

Tyler Black and Stan Kutcher. Suicide during covid-19: Myths, realities and lessons learned. UCBMJ, 2021.

Marta Blangiardo, Michela Cameletti, Monica Pirani, Gianni Corsetti, Marco Battaglini, and Gianluca Baio. Estimating weekly excess mortality at sub-national level in Italy during the COVID-19 pandemic. PloS One, 15(10):e0240286, 2020.

Debra M Boka and Howard Wainer. How can we estimate the death toll from COVID-19? CHANCE, 33(3): 67–72, 2020.

D Bradshaw, RE Dorrington, R Laubscher, TA Moultrie, and P Groenewald. Tracking mortality in near to real time provides essential information about the impact of the COVID-19 pandemic in South Africa in 2020. South African Medical Journal, 2021.

Renzo JC Calderon-Anyosa and Jay S Kaufman. Impact of COVID-19 lockdown policy on homicide, suicide, and motor vehicle deaths in Peru. Preventive Medicine, 143:106331, 2020.

Bimandra A Djaafara, Charles Whittaker, Oliver J Watson, Robert Verity, Nicholas F Brazeau, Widyastuti Widyastuti, Dwi Oktavia, Verry Adrian, Ngabila Salama, Sangeeta Bhatia, et al. Quantifying the dynamics of COVID-19 burden and impact of interventions in Java, Indonesia. medRxiv, 2020.

Ensheng Dong, Hongru Du, and Lauren Gardner. An interactive web-based dashboard to track COVID-19 in real time. The Lancet Infectious Diseases, 20(5):533–534, 2020.

European Commission. Road safety: 4000 fewer people lost their lives on eu roads in 2020 as death rate falls to all time low. Technical report, European Commission, 2021. URL https://ec.europa.eu/transport/modes/road/news/2021-04-20-road-safety_en.

William Farr. Vital statistics: a memorial volume of selections from the reports and writings of William Farr. Offices of the Sanitary Institute, London, 1885.

CP Farrington, Nick J Andrews, AD Beale, and MA Catchpole. A statistical algorithm for the early detection of outbreaks of infectious disease. Journal of the Royal Statistical Society: Series A (Statistics in Society), 159 (3):547–563, 1996.

Jeremy S Faust, Chengan Du, Katherine Dickerson Mayes, Shu-Xia Li, Zhenqiu Lin, Michael L Barnett, and Harlan M Krumholz. Mortality from drug overdoses, homicides, unintentional injuries, motor vehicle crashes, and suicides during the pandemic, March–August 2020. JAMA, 2021.

Jeremy Samuel Faust, Zhenqiu Lin, and Carlos Del Rio. Comparison of estimated excess deaths in new york city during the COVID-19 and 1918 influenza pandemics. JAMA Network Open, 3(8):e2017527–e2017527, 2020.

Federal Criminal Police Office. Police crime statistics 2020. Technical report, 2021. URL https://www.bka.de/EN/CurrentInformation/PoliceCrimeStatistics/2020/pcs2020.

A Flynn, C McGreevy, and EC Mulkerrin. Why do older patients die in a heatwave? QJM: An International Journal of Medicine, 98(3):227–229, 2005.

Anne Fouillet, Grégoire Rey, Françoise Laurent, Gérard Pavillon, Stéphanie Bellec, Chantal Guihenneuc-Jouyaux, Jacqueline Clavel, Eric Jougla, and Denis Hémon. Excess mortality related to the August 2003 heat wave in France. International Archives of Occupational and Environmental Health, 80(1):16–24, 2006.

Anne Fouillet, Grégoire Rey, Vérène Wagner, Karine Laaidi, Pascal Empereur-Bissonnet, Alain Le Tertre, Philippe Frayssinet, Pierre Bessemoulin, Françoise Laurent, Perrine De Crouy-Chanel, et al. Has the impact of heat waves on mortality changed in France since the European heat wave of summer 2003? A study of the 2006 heat wave. International Journal of Epidemiology, 37(2):309–317, 2008.

Mahan Ghafari, Alireza Kadivar, and Aris Katzourakis. Excess deaths associated with the iranian COVID-19 epidemic: a province-level analysis. medRxiv, 2020.

Charlie Giattino, Hanna Ritchie, Max Roser, Ortiz-Ospina Esteban, and Hasell Joe. Excess mortality during the Coronavirus pandemic (COVID-19). Technical report, Our World in Data, 2020. URL https://ourworldindata.org/excess-mortality-covid.

Thomas Hale, Noam Angrist, Emily Cameron-Blake, Laura Hallas, Betriz Kira, Saptarshi Majumdar, Anna Petherick, Toby Phillips, Helen Tatlow, and Samuel Webster. Variation in government responses to COVID-19. BSG Working Paper Series. Blavatnik School of Government. University of Oxford, 2020.

Jere Housworth and Alexander D Langmuir. Excess mortality from epidemic influenza, 1957–1966. American Journal of Epidemiology, 100(1):40–48, 1974.

Nazrul Islam, Vladimir M Shkolnikov, Rolando J Acosta, Ilya Klimkin, Ichiro Kawachi, Rafael A Irizarry, Gianfranco Alicandro, Kamlesh Khunti, Tom Yates, Dmitri A Jdanov, Martin White, Sarah Lewington, and Ben Lacey. Excess deaths associated with covid-19 pandemic in 2020: age and sex disaggregated time series analysis in 29 high income countries. BMJ, 373:n1137, 2021.

Dmitry Kobak. Excess mortality reveals Covid’s true toll in Russia. Significance, 18(1):16, 2021.

Vasilis Kontis, James E Bennett, Theo Rashid, Robbie M Parks, Jonathan Pearson-Stuttard, Michel Guillot, Perviz Asaria, Bin Zhou, Marco Battaglini, Gianni Corsetti, et al. Magnitude, demographics and dynamics of the effect of the first wave of the COVID-19 pandemic on all-cause mortality in 21 industrialized countries. Nature Medicine, pages 1–10, 2020.

Stacey Kung, Marjan Doppen, Melissa Black, Tom Hills, and Nethmi Kearns. Reduced mortality in New Zealand during the COVID-19 pandemic. The Lancet, 2020.

Junko Kurita, Tamie Sugawara, Yoshiyuki Sugishita, and Yasushi Ohkusa. Excess mortality in suicide caused by covid-19 in japan. medRxiv, 2021. doi: 10.1101/2021.02.13.21251670. URL https://www.medrxiv.org/content/early/2021/04/12/2021.02.13.21251670.

Alexander D Langmuir. William Farr: founder of modern concepts of surveillance. International Journal of Epidemiology, 5(1):13–18, 1976.

David A Leon, Vladimir M Shkolnikov, Liam Smeeth, Per Magnus, Markéta Pechholdová, and Christopher I Jarvis. COVID-19: a need for real-time monitoring of weekly excess deaths. The Lancet, 395(10234):e81, 2020.

Andrew T Levin, William P Hanage, Nana Owusu-Boaitey, Kensington B Cochran, Seamus P Walsh, and Gideon Meyerowitz-Katz. Assessing the age specificity of infection fatality rates for COVID-19: systematic review, meta-analysis, and public policy implications. European Journal of Epidemiology, pages 1–16, 2020.

Peter Lloyd-Sherlock, Shah Ebrahim, Ramón Martínez, Martin McKee, Enrique Acosta, and Lucas Sempé. Estimation of All-Cause Excess Mortality by Age-Specific Mortality Patterns of COVID-19 Pandemic in Peru in 2020. SSRN Scholarly Paper, (ID 3820553), April 2021. URL https://papers.ssrn.com/abstract=3820553.

Milken Institute. Ascertainment of the estimated excess mortality from hurricane Maria in Puerto Rico. Technical report, Milken Institute School of Public Health, 2018. URL https://publichealth.gwu.edu/sites/default/files/downloads/projects/PRstudy/Acertainment%20of%20the%20Estimated%20Excess%20Mortality%20from%20Hurricane%20Maria%20in%20Puerto%20Rico.pdf.

Ministry of the Interior. Insecurity and delinquency in 2020: a first snapshot - interstats analysis no. 32. Technical report, 2021. URL https://www.interieur.gouv.fr/Interstats/Themes/Homicides/Insecurite-et-delinquance-en-2020-une-premiere-photographie-Interstats-Analyse-N-32.

Chirag Modi,Vanessa Böhm, Simone Ferraro, George Stein, and Uroš Seljak. Estimating COVID-19 mortality in Italy early in the COVID-19 pandemic. Nature communications, 12(1):1–9, 2021.

Christopher JL Murray, Alan D. Lopez, Brian Chin, Dennis Feehan, and Kenneth H. Hill. Estimation of potential global pandemic influenza mortality on the basis of vital registry data from the 1918–20 pandemic: a quantitative analysis. The Lancet, 368(9554):2211–2218, 2006.

Lawrence Mwananyanda, Christopher J Gill, William MacLeod, Geoffrey Kwenda, Rachel Pieciak, Zachariah Mupila, Rotem Lapidot, Francis Mupeta, Leah Forman, Luunga Ziko, et al. Covid-19 deaths in Africa: prospective systematic postmortem surveillance study. BMJ, 372, 2021.

National Safety Council. Motor vehicle deaths in 2020 estimated to be highest in 13 years, despite dramatic drops in miles driven. Technical report, 2021. URL https://www.nsc.org/newsroom/motor-vehicle-deaths-2020-estimated-to-be-highest.

László Németh, Dmitri A Jdanov, and Vladimir M Shkolnikov. An open-sourced, web-based application to analyze weekly excess mortality based on the Short-term Mortality Fluctuations data series. PloS ONE, 16 (2):e0246663, 2021.

Angela Noufaily, Doyo G Enki, Paddy Farrington, Paul Garthwaite, Nick Andrews, and Andre Charlett. An improved algorithm for outbreak detection in multiple surveillance systems. Statistics in medicine, 32(7): 1206–1222, 2013.

Megan O’Driscoll, Gabriel Ribeiro Dos Santos, Lin Wang, Derek AT Cummings, Andrew S Azman, Juliette Paireau, Arnaud Fontanet, Simon Cauchemez, and Henrik Salje. Age-specific mortality and immunity patterns of SARS-CoV-2. Nature, 590(7844):140–145, 2021.

Eskild Petersen, Marion Koopmans, Unyeong Go, Davidson H Hamer, Nicola Petrosillo, Francesco Castelli, Merete Storgaard, Sulien Al Khalili, and Lone Simonsen. Comparing SARS-CoV-2 with SARS-CoV and influenza pandemics. The Lancet Infectious Diseases, 2020.

Jane Pirkis, Ann John, Sangsoo Shin, Marcos DelPozo-Banos, Vikas Arya, Pablo Analuisa-Aguilar, Louis Appleby, Ella Arensman, Jason Bantjes, Anna Baran, et al. Suicide trends in the early months of the COVID-19 pandemic: an interrupted time-series analysis of preliminary data from 21 countries. The Lancet Psychiatry, 0(0), 2021.

Tim Riffe and Enrique Acosta. Data resource profile: COVerAGE-DB: a global demographic database of COVID-19 cases and deaths. International Journal of Epidemiology, 50(2):390–390f, 2021.

Natalia Bustos Sierra, Nathalie Bossuyt, Toon Braeye, Mathias Leroy, Isabelle Moyersoen, Ilse Peeters, Aline Scohy, Johan Van der Heyden, Herman Van Oyen, and Françoise Renard. All-cause mortality supports the COVID-19 mortality in Belgium and comparison with major fatal events of the last century. Archives of Public Health, 78(1):1–8, 2020.

Lone Simonsen, Peter Spreeuwenberg, Roger Lustig, Robert J Taylor, Douglas M Fleming, Madelon Kroneman, Maria D Van Kerkhove, Anthony W Mounts, W John Paget, et al. Global mortality estimates for the 2009 influenza pandemic from the GLaMOR project: a modeling study. PLoS Medicine, 10(11):e1001558, 2013.

Thomas Spoorenberg. Data and methods for the production of national population estimates: An overview and analysis of available metadata. Technical report, United Nations, Department of Economics and Social Affairs, Population Division, 2020. URL https://www.un.org/development/desa/pd/content/data-and-methods-production-national-population-estimates-overview-and-analysis-available.

STMF. Short-term Mortality Fluctuation data series. Human Mortality Database. University of California Berkeley (USA) and Max Planck Institute for Demographic Research (Germany), 2021. https://www.mortality.org.

Takanao Tanaka and Shohei Okamoto. Increase in suicide following an initial decline during the COVID-19 pandemic in Japan. Nature Human Behaviour, 5(2):229–238, 2021.

The Economist. There have been 7m–13m excess deaths worldwide during the pandemic. Technical report, 2021. URL https://www.economist.com/briefing/2021/05/15/there-have-been-7m-13m-excess-deaths-worldwide-during-the-pandemic.

Transport Community. Fatalities for 2020 annual statistics for western balkans. Technical report, 2021. URL https://www.transport-community.org/wp-content/uploads/2021/04/Annual-Statistics-2020.pdf.

UNSD. Demographic yearbook 2019. Technical report, United Nations, Statistics Division, 2019. URL https://unstats.un.org/unsd/demographic-social/products/dyb/dyb_2019/.

Javier Vargas-Herrera, Karim Pardo Ruiz, Gladys Garro Nuñez Janet Miki Ohno, José Enrique Pérez-Lu, William Valdez Huarcaya, Benjamin Clapham, and Juan Cortez-Escalante. Resultados preliminares del fortalecimiento del sistema informático nacional de defunciones. Revista Peruana de Medicina Experimental y Salud Pública, 35:505–514, 2018.

Cécile Viboud, Rebecca F Grais, Bernard AP Lafont, Mark A Miller, and Lone Simonsen. Multinational impact of the 1968 Hong Kong influenza pandemic: evidence for a smoldering pandemic. The Journal of Infectious Diseases, 192(2):233–248, 2005.

Cécile Viboud, Lone Simonsen, Rodrigo Fuentes, Jose Flores, Mark A Miller, and Gerardo Chowell. Global mortality impact of the 1957–1959 influenza pandemic. The Journal of Infectious Diseases, 213(5):738–745, 2016.

Oliver J. Watson, Nada Abdelmagid, Aljaile Ahmed, Abd Elhameed Ahmed Abd Elhameed, Charles Whittaker, Nicholas Brazeau, Arran Hamlet, Patrick Walker, James Hay, Azra Ghani, Francesco Checchi, and Maysoon Dahab. Report 39 - Characterising COVID-19 epidemic dynamics and mortality under-ascertainment in Khartoum, Sudan. Technical report, MRC Centre for Global Infectious Disease Analysis, 2020. URL https://www.imperial.ac.uk/mrc-global-infectious-disease-analysis/covid-19/report-39-sudan.

Oliver J Watson, Mervat Alhaffar, Zaki Mehchy, Charles Whittaker, Zack Akil, Nicholas F Brazeau, Gina Cuomo-Dannenburg, Arran Hamlet, Hayley A Thompson, Marc Baguelin, et al. Leveraging community mortality indicators to infer COVID-19 mortality and transmission dynamics in Damascus, Syria. Nature Communications, 12(1):1–10, 2021.

Daniel M Weinberger, Jenny Chen, Ted Cohen, Forrest W Crawford, Farzad Mostashari, Don Olson, Virginia E Pitzer, Nicholas G Reich, Marcus Russi, Lone Simonsen, et al. Estimation of excess deaths associated with the COVID-19 pandemic in the United States, March to May 2020. JAMA Internal Medicine, 180(10):1336–1344, 2020.

Cory Welt and Andrew S. Bowen. Azerbaijan and Armenia: The Nagorno-Karabakh conflict (R46651). Technical report, Congressional Research Service, 2021. URL https://fas.org/sgp/crs/row/R46651.pdf.

Steven H Woolf, Derek A Chapman, Roy T Sabo, Daniel M Weinberger, and Latoya Hill. Excess deaths from COVID-19 and other causes, March-April 2020. JAMA, 324(5):510–513, 2020a.

Steven H Woolf, Derek A Chapman, Roy T Sabo, Daniel M Weinberger, Latoya Hill, and DaShaunda DH Taylor. Excess deaths from COVID-19 and other causes, March-July 2020. JAMA, 324(15):1562–1564, 2020b.

